# LioNeo project: a phase 2, randomized, double-blind, non-inferiority trial of a lyophilized human milk-based fortifier for very low birth weight infants

**DOI:** 10.64898/2025.12.10.25342013

**Authors:** Maria C. Achcar-Feih, Adriana Carnevale-da-Silva, Philippe P. Paiva, Tatiana Soares, Amanda Ansani, Vanessa S. Bomfim, Mariana M. Oliveira, Larissa G. Alves, Tânia Maria B. Trevilato, Isabela S. Dias, Marisa M. Mussi-Pinhata, Fábio Carmona, Davi C. Aragon, Fábio V. Ued, Luciana M. M. Fonseca, Vicky Nogueira-Pileggi, José S. Camelo

**Affiliations:** Department of Pediatrics, University of São Paulo, Ribeirão Preto, São Paulo, Brazil; Human Milk Bank, HC Criança – Clinics Hospital, University of São Paulo, Ribeirão Preto, São Paulo, Brazil; Laboratory of Metals and Rare Diseases / Pediatrics, Clinics Hospital, University of São Paulo, Ribeirão Preto, São Paulo, Brazil; Department of Health Sciences, Ribeirão Preto Medical School, University of São Paulo, Ribeirão Preto, São Paulo, Brazil; Department of Maternal-Infant and Public Health Nursing, University of São Paulo at Ribeirão Preto College of Nursing - WHO Collaborating Centre for Nursing Research Development, Ribeirão Preto, São Paulo, Brazil

**Author notes:** Corresponding author: José Simon Camelo Júnior (JSCJ) Department of Pediatrics, Ribeirão Preto Medical School, University of São Paulo. Avenida Bandeirantes, 3900, 5th floor, room 506, HC Criança – Clinics Hospital of Ribeirão Preto Medical School, University of São Paulo (HCFMRP-USP), Ribeirão Preto, São Paulo, Brazil. These authors contributed equally to this work (Maria C. Achcar-Feih, Adriana Carnevale-da-Silva, Vanessa S. Bomfim, Mariana M. Oliveira, Larissa G. Alves, Fábio V. Ued, Vicky Nogueira-Pileggi, José S. Camelo). These authors also contributed equally to this work (Philippe P. Paiva, Tatiana Soares, Amanda Ansani, Tânia Maria B. Trevilato, Isabela S. Dias, Marisa M. Mussi-Pinhata, Fábio Carmona, Davi C. Aragon, Luciana M. M. Fonseca).

**Keywords:** Freeze Drying, Infant, Premature, Milk Banks, Milk, Human, Randomized Controlled Trial

## Abstract

**Background:** Cow’s milk protein-based fortifiers are the standard for human milk fortification in very low birth weight (VLBW) infants but have been associated with a higher risk of necrotizing enterocolitis. LioNeo is produced by lyophilization of donor human milk. This study assessed whether LioNeo is non-inferior to the bovine fortifier for growth in VLBW infants.

**Methods:** This phase 2, randomized, double-blind, non-inferiority trial was conducted at a university hospital. Hemodynamically stable VLBW infants who were small or appropriate for gestational age and exclusively fed human milk (≥100 mL/kg/day) were randomized 1:1 to LioNeo or human milk with a bovine fortifier (HMCA) for 21 days. The primary outcomes were length and head circumference z-scores. Secondary outcomes were weight gain, adverse events, and serum copper and zinc concentrations.

**Results:** In total, 129 infants were randomized and analyzed by intention to treat (LioNeo, 67; HMCA, 62). At day 21, the adjusted difference in length z-score was 0.07 (95% CI −0.13 to 0.27), with the lower limit above the non-inferiority margin (−0.27). There was no difference in head circumference or weight. Late-onset neonatal sepsis occurred in six HMCA participants and none in the LioNeo group (*P* = 0.01). Serum zinc decreased in both groups; copper was higher with LioNeo at day 21.

**Conclusions:** LioNeo was non-inferior to the bovine fortifier for linear growth in VLBW infants, with no difference in head circumference or weight and no increase in adverse events. Multicenter phase 3 trials are needed.

**Trial registration:** Brazilian Registry of Clinical Trials (ReBEC), RBR-8nnpfm (UTN U1111-1220-0550).

## INTRODUCTION

Neonatal mortality accounts for most deaths among children under 5 years of age worldwide, predominantly due to preterm birth and its complications (1). In this context, adequate nutrition is a key determinant of outcomes in very low birth weight (VLBW) infants (birth weight ≤1,500 g). Human milk (HM) is recognized as the gold standard feed for VLBW infants because of its immunological support, digestibility, and nutritional composition. The World Health Organization (WHO) recommends donor HM when the mother’s own milk is not available (2). However, HM alone provides insufficient protein, minerals, and micronutrients to meet the growth demands of this population, making fortification mandatory (3).

Fortifiers based on hydrolyzed cow’s milk protein have established nutritional efficacy and represent the current standard of care. However, their use has been associated with a higher risk of necrotizing enterocolitis and sensitization to heterologous protein, whereas exclusive HM-based diets, with fortifiers derived from HM itself, have shown safety advantages (4,5). These fortifiers are produced commercially in the United States but cannot be used in Brazil and other countries that prohibit the commercialization of human fluids (6).

Israel-Ballard et al. issued a call to action for equitable access to HM for vulnerable infants (7). Providing an exclusive HM-based diet to VLBW infants requires an HM-derived fortifier that is feasible in public health systems. To meet this need, LioNeo was developed as a concentrate produced by lyophilization of donor HM, using a simplified method designed for implementation in human milk banks. Preclinical studies demonstrated physicochemical stability, microbiological safety, and a nutritional increase of approximately 60% relative to baseline HM (8). A phase 1 trial with 40 VLBW infants confirmed that LioNeo is safe and well tolerated in hemodynamically stable VLBW infants (9).

Given that the cow’s milk protein-based fortifier has established nutritional efficacy and represents the current standard of care for VLBW infants, and that LioNeo offers potential advantages in safety, feasibility in public health systems, and alignment with the recommendation for an exclusive HM-based diet, a non-inferiority trial was conducted to evaluate this intervention. In this design, if LioNeo demonstrates non-inferior linear growth compared with the current standard, its additional advantages would represent an important advance for this population.

This study aimed to assess whether LioNeo is non-inferior to the cow’s milk protein-based fortifier for linear growth (length) and head circumference in VLBW infants followed for 21 days, as well as for weight gain, adverse events, and serum copper and zinc concentrations.

## METHODS

### Study design and setting

This phase 2, randomized, controlled, double-blind, non-inferiority trial with 1:1 allocation was conducted in the neonatal intensive care and intermediate care units of the Clinics Hospital of Ribeirão Preto Medical School, University of São Paulo (HCFMRP-USP), Brazil, between October 2020 and August 2023. The protocol was approved by the Research Ethics Committee (CAAE 96682318.2.0000.5440) and registered under Universal Trial Number (UTN) U1111-1220-0550 (http://www.ensaiosclinicos.gov.br/rg/RBR-8nnpfm/) on June 4, 2019. Parents or legal guardians provided written informed consent (Supplementary File 1). The study followed the Declaration of Helsinki (10) and is reported in accordance with the CONSORT 2025 statement (11).

### Participants

Infants were eligible if they had a birth weight ≤1,500 g, gestational age <37 weeks, were small or appropriate for gestational age, were exclusively fed HM (≥100 mL/kg/day), and were hemodynamically stable (no vasoactive drugs, blood pressure within the normal range for birth weight and postnatal age and/or good peripheral perfusion, and oxygen saturation >90%). For monozygotic twins, only one infant was randomized. Exclusion criteria were large for gestational age, major malformations, grade III or higher peri- or intraventricular hemorrhage, moderate or severe perinatal asphyxia, previous necrotizing enterocolitis or feeding intolerance, and minor mothers without a legal guardian.

### Randomization and blinding

The allocation sequence was generated electronically by the study statistician on the Sealed Envelope platform using randomly sized blocks (four or six). Allocation concealment was ensured by REDCap (Research Electronic Data Capture), which revealed the allocation only at enrollment; the nurses responsible for recruitment had no access to the list. Parents, clinical staff, investigators, and outcome assessors were blinded. Only the three nutritionists responsible for preparing the feeds and the pharmacist who handled HM and LioNeo were unblinded. The feeds were indistinguishable in appearance, color, and odor, and groups were identified by codes on the syringes and in the database.

### Intervention and control

The intervention group received LioNeo, produced at the HCFMRP-USP human milk bank by reconstituting approximately 7 g of lyophilized HM (derived from 50 mL of donor milk) in 75 mL of fluid HM, followed by pasteurization and microbiological quality control. The control group received donor HM fortified with FM85® (Nestlé®), a hydrolyzed cow’s milk protein-based additive, at 4% (1 g per 25 mL), hereafter referred to as HMCA. The composition of the feeds is shown in Table I. Detailed descriptions of the development, testing, and production of LioNeo are available in the preclinical and phase 1 publications (8,9).

**Table I.** Energy, macronutrient and micronutrient content per 100 mL of baseline human milk, LioNeo and HMCA.

| Component (per 100 mL) | Baseline human milk | LioNeo | HMCA |
| --- | --- | --- | --- |
|  | Mean (sd) | Mean (sd) | Mean (sd) |
| Energy (kcal) | 56.3 (10.5) | 80.0 (13.7) | 73.5 (10.5) |
| Protein (g) | 0.9 (0.5) | 1.5 (0.6) | 2.3 (0.5) |
| Carbohydrates (g) | 7.1 (0.7) | 9.2 (0.7) | 8.4 (0.7) |
| Total lipids (g) | 2.6 (1.1) | 4.0 (1.4) | 3.3 (1.1) |
| Calcium (mg) | 23.2 (4.7) | 36.5 (7.2) | 99.2 (4.7) |
| Phosphorus (mg) | 14.8 (6.0) | 18.5 (6.0) | 58.5 (6.3) |
| Magnesium (mg) | 2.3 (0.5) | 3.7 (0.7) | 6.3 (0.5) |
| Sodium (mg) | 13.1 (9.8) | 22.3 (16.9) | 49.9 (9.1) |
| Potassium (mg) | 60.1 (14.8) | 101.4 (26.9) | 108.1 (14.8) |
| Iron (mg) | 0.1 (0.0) | 0.1 (0.0) | 1.9 (0.0) |
| Zinc (mg) | 0.1 (0.1) | 0.2 (0.1) | 1.1 (0.1) |
| Copper (µg) | 33.4 (14.6) | 48.3 (19.9) | 85.4 (14.6) |
| Protein:energy ratio (g/100 kcal)<sup>a</sup> | 1.60 | 1.88 | 3.13 |

sd: standard deviation. Baseline human milk: pasteurized donor human milk without fortification; LioNeo: human milk concentrated with human milk lyophilizate; HMCA: human milk with commercial additive based on cow's milk protein (FM85®, Nestlé®, 4%). <sup>a</sup>Calculated from mean protein and energy values. Adapted from Nogueira-Pileggi et al. (9).

Feeds were administered according to daily medical prescriptions and the unit’s enteral nutrition protocol (100 to 160 mL/kg/day). The 21-day follow-up period was defined by consensus of the research team, based on similar nutritional intervention studies in VLBW infants (12). Decisions on interrupting or modifying feeds were made by the clinical team, without interference from the research team. Expressed mother’s own milk was prioritized and, when available, the study feed was offered as a complement. Daily assessments included body weight, abdominal circumference, volume of milk ingested, and adverse events, recorded in REDCap by trained nurses and double-checked by the project manager. The enteral nutrition protocol and the standard operating manuals for feed portioning and data collection are provided in Supplementary Files 2, 3, and 4, respectively.

### Outcomes

The primary outcomes were linear growth, assessed by length, and head circumference, measured on the first (day 1) and twenty-first (day 21) study days with an acrylic stadiometer adaptable to the incubator and a non-stretchable measuring tape, respectively (precision 0.1 cm), and converted into z-scores using the INTERGROWTH-21st standards (13). Secondary outcomes were weight gain (Filizola Baby® electronic scale, precision 5 g), adverse events, and serum copper and zinc concentrations on day 1 and day 21, measured by atomic absorption spectrophotometry (Varian®, Mulgrave, Australia).

Predefined adverse events were diarrhea, regurgitation, vomiting, abdominal distension, gastrointestinal bleeding, gastrointestinal perforation, necrotizing enterocolitis (Bell stage IB or higher), late-onset neonatal sepsis (suspected or culture-confirmed), and death, monitored daily by a physician from the research team (Supplementary Table S1). All serious adverse events were reported to an independent external safety committee, which also conducted an interim analysis at the study midpoint. The stopping criterion was a significantly higher frequency of serious events in the intervention group, based on Vermont Oxford Network frequencies (14).

### Sample size

The sample size was calculated for length, with no specific margin for head circumference. The non-inferiority margin corresponded to 11% of the expected difference between group means in length (0.97 cm), converted to a z-score using a standard deviation of 0.40, resulting in 0.27 [(0.11 × 0.97) / 0.40]. With 80% power and a 5% significance level, the calculated sample size was 160 VLBW infants per group, totaling 320 participants. Owing to recruitment difficulties during the COVID-19 pandemic, recruitment ended in August 2023, and the analysis was conducted with the sample obtained, maintaining the prespecified non-inferiority criterion.

### Statistical analysis

Group means on day 1 and day 21 were compared using linear mixed-effects models, simple and adjusted for cumulative milk intake and baseline values (day 1), with differences between means estimated by orthogonal contrasts and two-sided 95% confidence intervals (15). Cumulative milk intake was included as a covariate because it represents exposure to the intervention, recorded daily. For adverse events, participants with at least one occurrence during the 21 days were considered positive; relative risks were estimated by log-binomial regression, simple and adjusted for cumulative milk intake, and Fisher’s exact test was used when there were zero events in one group. Event counts per participant were compared using double Poisson models, simple and adjusted.

Analyses were performed by intention to treat (ITT), including all randomized participants according to their allocated group, and per protocol (PP), including participants with primary outcome measurements on day 1 and day 21, as recommended for non-inferiority trials (16,17). The mixed-effects models used all available observations, without imputation, under the missing at random (MAR) assumption. Analyses were performed using SAS 9.4 (SAS Institute, Cary, NC, USA) and R 4.5.0.

### AI-assisted writing

We used Claude (Anthropic) to support translation from Portuguese into English, text editing, and language refinement during the preparation of this manuscript. The tool was used to improve English language clarity, as the authors are non-native English speakers. All AI-generated suggestions were carefully reviewed and edited by the authors, who take full responsibility for the final content.

## RESULTS

Between October 15, 2020, and August 9, 2023, 201 newborns were assessed for eligibility; 72 were excluded and 129 were randomized (LioNeo, 67; HMCA, 62). All received the allocated intervention and were included in the ITT analysis. On day 21, primary outcome data were available for 61 participants in the LioNeo group and 55 in the HMCA group, who made up the PP analysis. The flow of participants and the reasons for exclusion and missing data are shown in Fig. 1.

**Fig. 1.**
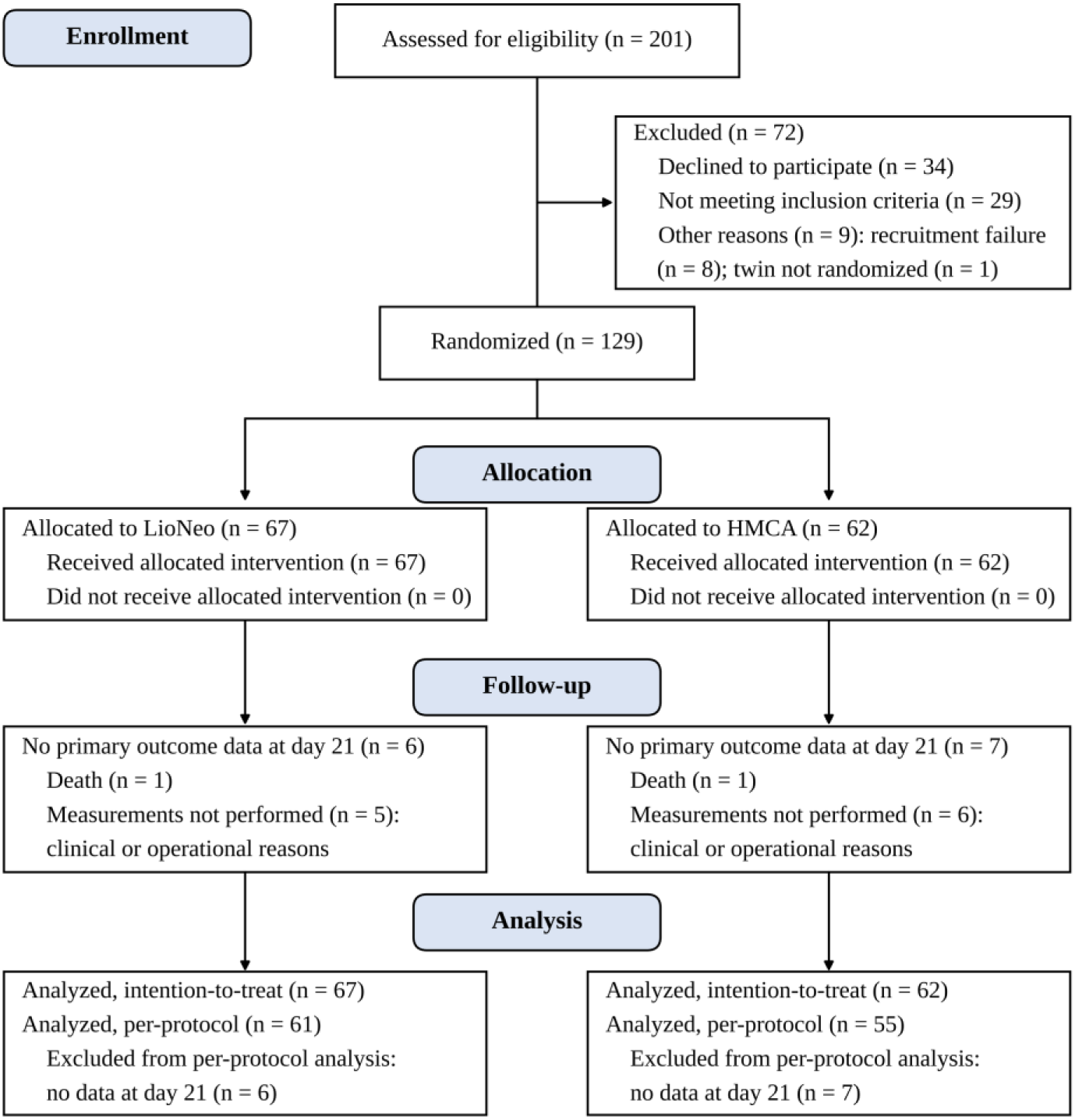
Flow diagram of participants Adapted from the CONSORT 2025 statement (11). LioNeo: human milk concentrated with human milk lyophilizate; HMCA: human milk with commercial additive based on cow’s milk protein.

Characteristics at birth and at enrollment were similar between groups (Table II), with a slight predominance of females (LioNeo, 38/67, 56.7%; HMCA, 32/62, 51.6%). Anthropometric measurements, milk intake, and serum copper and zinc concentrations are shown in Table III. Day 1 anthropometric measurements refer to all randomized participants (ITT) and were similar between groups; day 21 measurements refer to participants assessed on that day (PP). Mean daily milk intake was similar between groups (188.40 and 187.61 mL), and the mean number of days on the study feed was 17.75 in the LioNeo group and 15.45 in the HMCA group.

**Table II.**
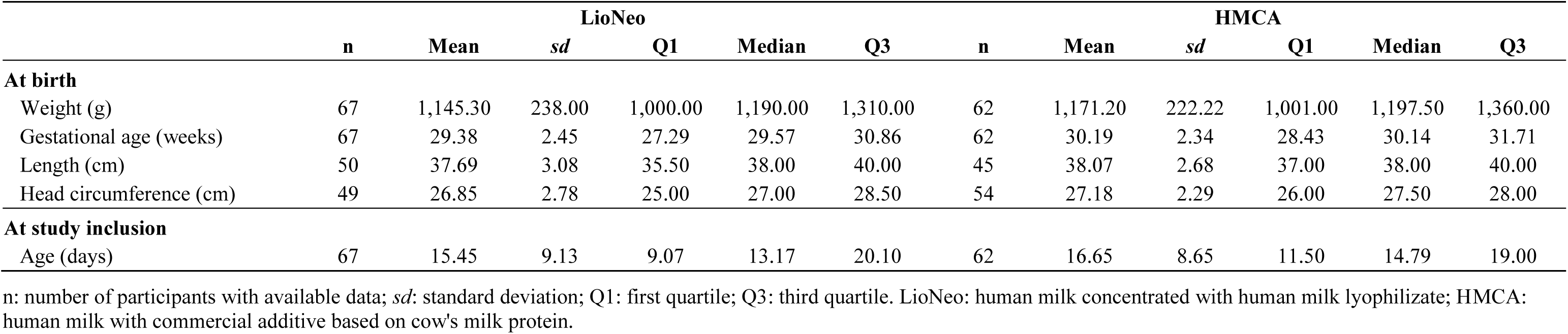
Characteristics of the preterm infants at birth and at study inclusion by group.

**Table III.** Anthropometric measurements, milk intake and serum minerals at day 1 and day 21 by group.

|  | LioNeo |  |  |  |  |  | HMCA |  |  |  |  |  |
| --- | --- | --- | --- | --- | --- | --- | --- | --- | --- | --- | --- | --- |
|  | n | Mean | sd | Q1 | Median | Q3 | n | Mean | sd | Q1 | Median | Q3 |
| **Day 1 (ITT)** |  |  |  |  |  |  |  |  |  |  |  |  |
| Weight (g) | 67 | 1,194.03 | 233.51 | 1,000.00 | 1,240.00 | 1,370.00 | 62 | 1,253.47 | 203.52 | 1,110.00 | 1,295.00 | 1,400.00 |
| Weight z-score | 67 | -1.54 | 1.04 | -2.24 | -1.53 | -0.76 | 62 | -1.77 | 1.05 | -2.66 | -1.76 | -1.09 |
| Length (cm) | 67 | 38.78 | 3.32 | 36.00 | 39.00 | 40.00 | 62 | 39.18 | 3.56 | 38.00 | 39.00 | 41.00 |
| Length z-score | 67 | -1.17 | 1.44 | -2.31 | -1.13 | -0.21 | 62 | -1.54 | 1.63 | -2.65 | -1.51 | -0.54 |
| Head circumference (cm) | 67 | 26.49 | 2.15 | 25.00 | 26.70 | 28.00 | 62 | 26.75 | 2.01 | 25.60 | 27.00 | 28.00 |
| Head circumference z-score | 67 | -1.66 | 1.24 | -2.46 | -1.71 | -0.91 | 62 | -2.05 | 1.27 | -2.93 | -2.21 | -1.25 |
| **Day 21 (PP)** |  |  |  |  |  |  |  |  |  |  |  |  |
| Weight (g) | 61 | 1,560.52 | 320.58 | 1,310.00 | 1,625.00 | 1,795.00 | 55 | 1,609.27 | 271.85 | 1,390.00 | 1,590.00 | 1,840.00 |
| Weight z-score | 61 | -1.92 | 0.90 | -2.44 | -1.98 | -1.38 | 55 | -2.17 | 0.94 | -2.95 | -2.13 | -1.57 |
| Length (cm) | 61 | 41.12 | 3.02 | 39.00 | 42.00 | 43.00 | 55 | 41.06 | 3.76 | 39.70 | 42.00 | 43.00 |
| Length z-score | 61 | -1.81 | 1.43 | -2.72 | -2.04 | -0.98 | 55 | -2.23 | 1.59 | -3.39 | -2.14 | -1.17 |
| Head circumference (cm) | 61 | 28.08 | 2.34 | 26.00 | 28.00 | 30.00 | 55 | 28.42 | 1.85 | 27.50 | 28.50 | 30.00 |
| Head circumference z-score | 61 | -2.22 | 1.19 | -3.00 | -2.28 | -1.47 | 55 | -2.53 | 1.08 | -3.33 | -2.63 | -1.83 |
| **Milk intake** |  |  |  |  |  |  |  |  |  |  |  |  |
| Total milk intake (mL) | 67 | 3,445.96 | 1,453.90 | 2,480.00 | 3,668.00 | 4,639.00 | 62 | 3,108.00 | 1,705.72 | 2,020.00 | 3,604.00 | 4,541.00 |
| Days of milk intake | 67 | 17.75 | 5.88 | 17.00 | 21.00 | 21.00 | 62 | 15.45 | 7.32 | 12.00 | 21.00 | 21.00 |
| Mean daily milk intake (mL) | 67 | 188.40 | 45.50 | 159.30 | 192.38 | 223.60 | 62 | 187.61 | 45.45 | 162.50 | 192.18 | 221.00 |
| **Serum minerals (µg/dL)** |  |  |  |  |  |  |  |  |  |  |  |  |
| Copper, day 1 | 60 | 54.15 | 18.06 | 43.20 | 51.40 | 61.80 | 57 | 53.84 | 19.96 | 41.00 | 49.40 | 60.40 |
| Copper, day 21 | 49 | 61.06 | 18.24 | 46.20 | 60.20 | 67.80 | 50 | 43.18 | 11.85 | 35.40 | 42.60 | 51.00 |
| Zinc, day 1 | 60 | 80.17 | 16.78 | 71.50 | 81.55 | 89.50 | 58 | 76.71 | 15.99 | 65.50 | 77.50 | 85.50 |
| Zinc, day 21 | 50 | 66.08 | 13.42 | 59.00 | 68.00 | 76.50 | 50 | 61.57 | 14.36 | 52.00 | 59.50 | 71.00 |

ITT: intention-to-treat; PP: per-protocol; n: number of participants with available data; sd: standard deviation; Q1: first quartile; Q3: third quartile. LioNeo: human milk concentrated with human milk lyophilizate; HMCA: human milk with commercial additive based on cow's milk protein.

On day 21, the difference between means in length z-score (LioNeo − HMCA) was 0.07 (95% CI −0.13 to 0.27) in the adjusted model and 0.41 (95% CI −0.12 to 0.96) in the simple model; in the PP analysis, it was 0.06 (95% CI −0.14 to 0.27) and 0.42 (95% CI −0.12 to 0.96), respectively. In all approaches, the lower limit was above the non-inferiority margin of −0.27 (Table IV and Fig. 2). For head circumference z-score, there was no difference between groups: 0.31 (95% CI −0.16 to 0.70) in the simple model and −0.09 (95% CI −0.29 to 0.11) in the adjusted model, with similar results in the PP analysis. There was also no difference in weight z-score (adjusted difference 0.005; 95% CI −0.08 to 0.09). Between day 1 and day 21, both groups showed increases in weight, length, and head circumference (Table III), with a similar decrease in the respective z-scores (Table IV).

**Fig. 2.**
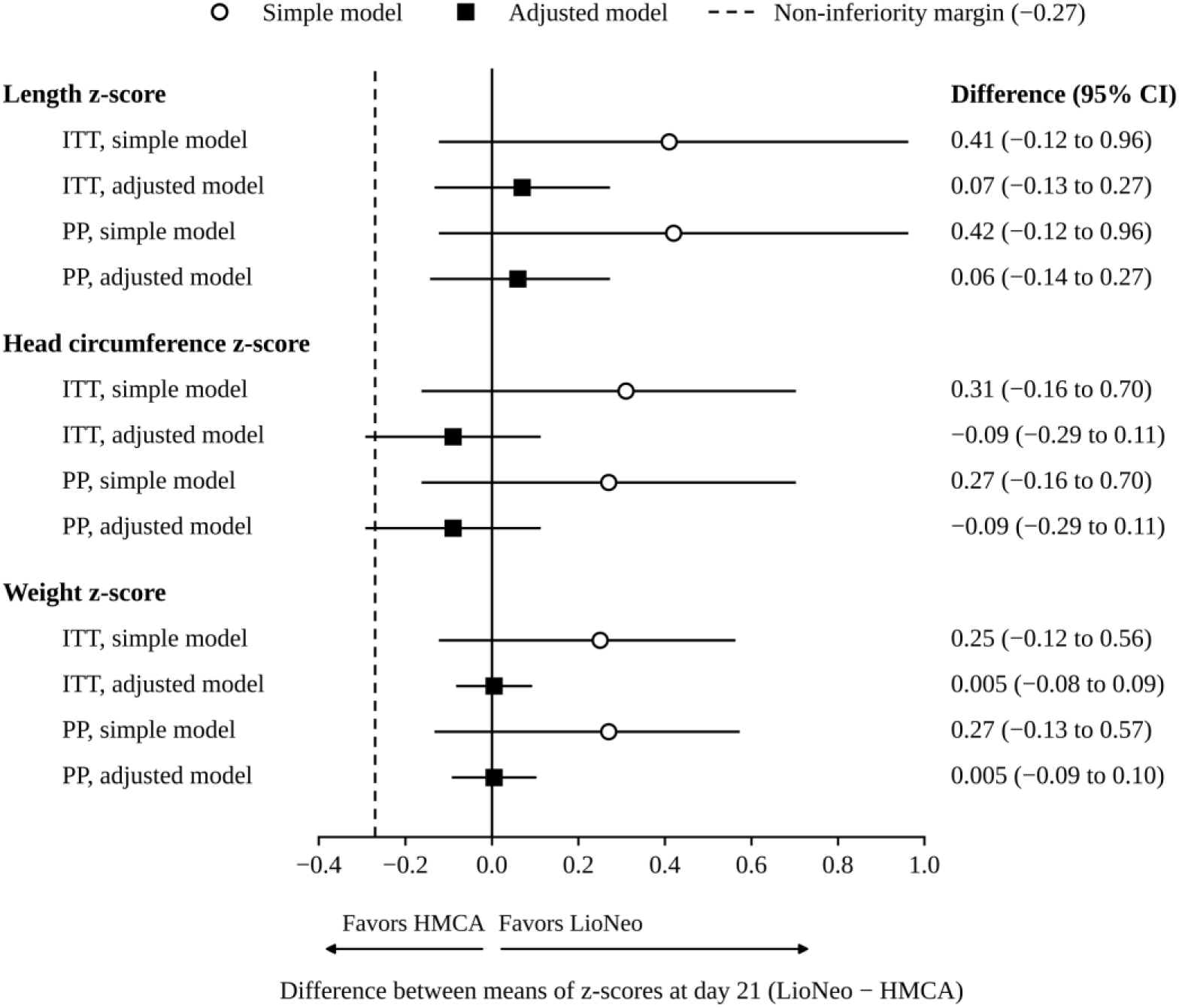
Differences in anthropometric z-scores at day 21: intention-to-treat and per-protocol analyses Differences between means (LioNeo − HMCA) and 95% confidence intervals estimated by linear mixed-effects models. Open circles: simple model; filled squares: model adjusted for cumulative milk intake and baseline values (day 1). The dashed vertical line indicates the non-inferiority margin (−0.27), prespecified for length only. ITT: intention-to-treat; PP: per-protocol. LioNeo: human milk concentrated with human milk lyophilizate; HMCA: human milk with commercial additive based on cow’s milk protein.

**Table IV.** Differences between means of anthropometric z-scores and serum minerals estimated by linear mixed-effects models.

| Comparison | Intention-to-treat |  | Per-protocol |  |
| --- | --- | --- | --- | --- |
|  | Simple model | Adjusted model | Simple model | Adjusted model |
| **Length z-score** |  |  |  |  |
| LioNeo – HMCA, day 1 | 0.37 (–0.16 to 0.90) | – | 0.37 (–0.15 to 0.90) | – |
| LioNeo – HMCA, day 21 | 0.41 (–0.12 to 0.96)<sup>a</sup> | 0.07 (–0.13 to 0.27)<sup>a</sup> | 0.42 (–0.12 to 0.96)<sup>a</sup> | 0.06 (–0.14 to 0.27)<sup>a</sup> |
| Day 21 – day 1, LioNeo | –0.66 (–0.88 to –0.46) | – | –0.67 (–0.88 to –0.46) | – |
| Day 21 – day 1, HMCA | –0.71 (–0.93 to –0.50) | – | –0.71 (–0.94 to –0.50) | – |
| **Head circumference z-score** |  |  |  |  |
| LioNeo – HMCA, day 1 | 0.39 (–0.02 to 0.81) | – | 0.40 (–0.03 to 0.82) | – |
| LioNeo – HMCA, day 21 | 0.31 (–0.16 to 0.70) | –0.09 (–0.29 to 0.11) | 0.27 (–0.16 to 0.70) | –0.09 (–0.29 to 0.11) |
| Day 21 – day 1, LioNeo | –0.57 (–0.79 to –0.36) | – | –0.58 (–0.79 to –0.36) | – |
| Day 21 – day 1, HMCA | –0.44 (–0.66 to –0.22) | – | –0.45 (–0.66 to –0.22) | – |
| **Weight z-score** |  |  |  |  |
| LioNeo – HMCA, day 1 | 0.23 (–0.11 to 0.58) | – | 0.23 (–0.11 to 0.58) | – |
| LioNeo – HMCA, day 21 | 0.25 (–0.12 to 0.56) | 0.005 (–0.08 to 0.09) | 0.27 (–0.13 to 0.57) | 0.005 (–0.09 to 0.10) |
| Day 21 – day 1, LioNeo | –0.40 (–0.52 to –0.32) | – | –0.42 (–0.52 to –0.33) | – |
| Day 21 – day 1, HMCA | –0.38 (–0.51 to –0.30) | – | –0.41 (–0.51 to –0.30) | – |
| **Copper (µg/dL)** |  |  |  |  |
| LioNeo – HMCA, day 1 | 0.50 (–5.87 to 6.89) | – | NA | NA |
| LioNeo – HMCA, day 21 | 18.26 (11.33 to 25.20) | 16.83 (11.49 to 22.19) | NA | NA |
| Day 21 – day 1, LioNeo | 6.72 (0.51 to 12.92) | – | NA | NA |
| Day 21 – day 1, HMCA | –11.03 (–17.36 to –4.72) | – | NA | NA |
| **Zinc (µg/dL)** |  |  |  |  |
| LioNeo – HMCA, day 1 | 3.51 (–2.08 to 9.11) | – | NA | NA |
| LioNeo – HMCA, day 21 | 3.38 (–2.68 to 9.43) | –0.35 (–4.89 to 4.20) | NA | NA |
| Day 21 – day 1, LioNeo | –14.77 (–19.93 to –9.61) | – | NA | NA |
| Day 21 – day 1, HMCA | –14.64 (–19.90 to –9.37) | – | NA | NA |

Data are differences between means (95% confidence interval). LioNeo: human milk concentrated with human milk lyophilizate; HMCA: human milk with commercial additive based on cow's milk protein. Adjusted model: adjusted for cumulative milk intake and baseline values (day 1). –: not applicable, as covariate adjustment is only possible at the end of the study. NA: not analyzed in the per-protocol analysis. <sup>a</sup>Lower limit of the 95% confidence interval above the prespecified non-inferiority margin for length (–0.27).

Serum copper was similar between groups on day 1 and higher in the LioNeo group on day 21 (61.06 versus 43.18 µg/dL; adjusted difference 16.83 µg/dL; 95% CI 11.49 to 22.19), increasing in the LioNeo group and decreasing in the HMCA group over follow-up. Serum zinc decreased similarly in both groups, with no difference between them on day 1 or day 21 (adjusted difference on day 21: −0.35 µg/dL; 95% CI −4.89 to 4.20) (Tables III and IV; Fig. 3).

**Fig. 3.**
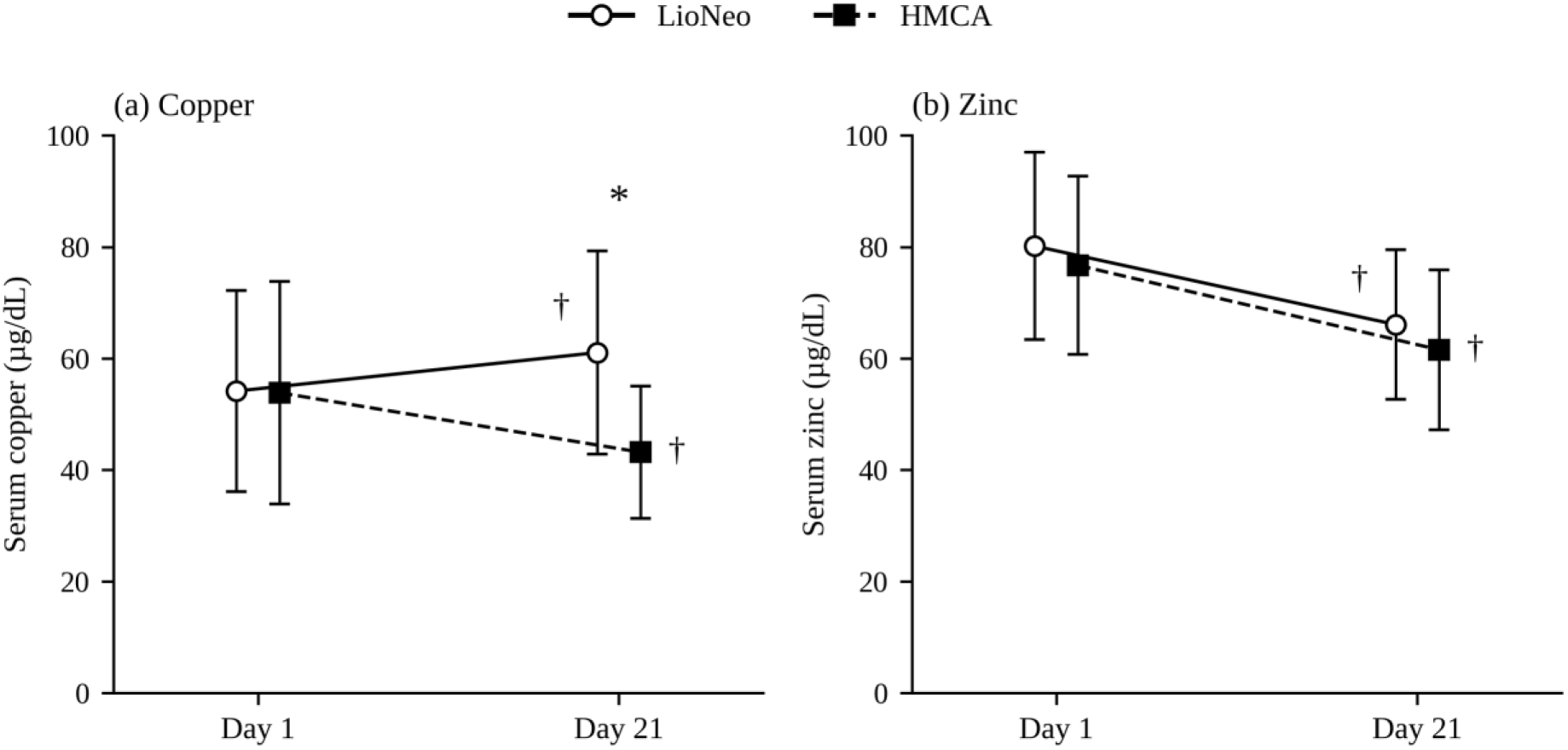
Serum copper and zinc concentrations at day 1 and day 21 by group Data are presented as mean ± standard deviation (values in Table III). Differences were estimated by linear mixed-effects models (Table IV). *Significant difference between groups at day 21. †Significant change from day 1 to day 21 within the group. LioNeo: human milk concentrated with human milk lyophilizate; HMCA: human milk with commercial additive based on cow’s milk protein.

Vomiting and abdominal distension were the most frequent adverse events (LioNeo: 29.85% and 23.88%; HMCA: 46.77% and 37.10%), with no difference between groups (Table V); for vomiting, the upper limit of the CI reached 1.00 in the simple model (RR 0.63; 95% CI 0.40 to 1.00) but not in the adjusted model (RR 0.68; 95% CI 0.43 to 1.08). Necrotizing enterocolitis occurred in one participant in the LioNeo group (1.49%) and in three in the HMCA group (4.84%) (adjusted RR 0.39; 95% CI 0.04 to 3.69), and there were no cases of diarrhea or gastrointestinal perforation. Late-onset neonatal sepsis occurred in six participants in the HMCA group (9.68%) and in none in the LioNeo group (*P* = 0.01; Fisher’s exact test), a result consistent with the PP analysis (five cases, 9.09%, versus none; *P* = 0.02) (Supplementary Table S2). In the comparison of event counts per participant using double Poisson models, necrotizing enterocolitis was lower in the LioNeo group in the simple model (difference −0.03; 95% CI −0.06 to −0.005) but not in the adjusted model (−0.10; 95% CI −0.19 to 0.004); the other events did not differ between groups (Supplementary Table S3). Two deaths occurred, one in each group; the independent external safety committee reviewed the cases with unblinded data, concluded that they were unrelated to the interventions, and deemed the study fit to continue.

**Table V.** Incidence of adverse events by group: intention-to-treat analysis.

| Adverse event | LioNeo (n = 67)<br>n (%) | HMCA (n = 62)<br>n (%) | RR (95% CI) | Adjusted RR (95% CI) |
| --- | --- | --- | --- | --- |
| Diarrhoea | 0 (0.00) | 0 (0.00) | —<sup>a</sup> | —<sup>a</sup> |
| Regurgitation | 2 (2.99) | 5 (8.06) | 0.37 (0.07 to 1.83) | 0.38 (0.08 to 1.90) |
| Vomiting | 20 (29.85) | 29 (46.77) | 0.63 (0.40 to 1.00) | 0.68 (0.43 to 1.08) |
| Abdominal distension | 16 (23.88) | 23 (37.10) | 0.64 (0.38 to 1.10) | 0.70 (0.41 to 1.19) |
| Gastrointestinal bleeding | 4 (5.97) | 6 (9.68) | 0.62 (0.18 to 2.08) | 0.63 (0.18 to 2.13) |
| Gastrointestinal perforation | 0 (0.00) | 0 (0.00) | —<sup>a</sup> | —<sup>a</sup> |
| Necrotizing enterocolitis | 1 (1.49) | 3 (4.84) | 0.30 (0.03 to 2.89) | 0.39 (0.04 to 3.69) |
| Late-onset neonatal sepsis | 0 (0.00) | 6 (9.68) | P = 0.01<sup>b</sup> | —<sup>a</sup> |

Incidence: participants with at least one occurrence of the event during the 21 days of study. RR: relative risk (LioNeo vs. HMCA), estimated by log-binomial regression; adjusted RR: adjusted for cumulative milk intake; CI: confidence interval. LioNeo: human milk concentrated with human milk lyophilizate; HMCA: human milk with commercial additive based on cow's milk protein. <sup>a</sup>Not estimable due to zero events in one or both groups. <sup>b</sup>Fisher's exact test.

## DISCUSSION

This phase 2 trial demonstrated that LioNeo is non-inferior to HMCA for linear growth in VLBW infants. In the ITT analysis, in the model adjusted for milk intake and baseline values, the estimated between-group difference in length z-score on day 21 was 0.07 (95% CI −0.13 to 0.27), with the lower limit above the prespecified non-inferiority margin of −0.27. This result was confirmed by the PP analysis (difference 0.06; 95% CI −0.14 to 0.27) and by the simple models, in which the non-inferiority criterion was also met, reinforcing its robustness. For head circumference, there was no difference between groups; using the margin defined for length as a reference, the lower limit of the confidence interval was above it in the simple model and slightly below it in the adjusted model (−0.29). Weight gain was similar between groups, no adverse event was more frequent in the LioNeo group, late-onset neonatal sepsis occurred only in the HMCA group, and serum copper concentrations were higher in the LioNeo group at the end of follow-up.

The similar changes in length, head circumference, and weight between groups are relevant given that LioNeo provides less protein than HMCA (1.5 versus 2.3 g/100 mL) and more energy (80.0 versus 73.5 kcal/100 mL), with protein: energy ratios of 1.88 and 3.13 g/100 kcal, respectively (Table I). These findings suggest that growth in VLBW infants depends on the overall nutritional balance of the diet rather than on protein concentration alone, in line with evidence that HM-based fortification strategies can support adequate growth even when protein intake falls below traditionally recommended values (18,19). Lavassani et al. also observed equivalent gains in weight, length, and head circumference with human or bovine fortifiers (20). The similar decrease in z-scores in both groups, despite increases in absolute measurements, reflects the growth deceleration commonly observed in VLBW infants in the first weeks of life, for which ESPGHAN recommends monitoring individual growth trajectories (3); because it was similar between groups, it does not affect the comparison between fortifiers.

Follow-up studies suggest that exclusive HM-based diets may benefit body composition and neurodevelopment, outcomes not assessed in the present study (21,22). In a systematic review, Suganuma et al. considered the evidence on HM and short-term growth in preterm infants inconclusive and noted that only one study with an HM-derived fortifier had assessed growth (23). In this context, the present trial adds growth data obtained with an HM-derived fortifier, which supported growth similar to that achieved with the bovine fortifier.

Vomiting, regurgitation, abdominal distension, and gastrointestinal bleeding were numerically less frequent in the LioNeo group, without statistically significant differences, consistent with the phase 1 safety data (9). Necrotizing enterocolitis occurred in one participant in the LioNeo group and in three in the HMCA group, with no difference in incidence (adjusted RR 0.39; 95% CI 0.04 to 3.69); in the comparison of counts per participant, the difference was significant in the simple model but not in the adjusted model (Supplementary Table S3). The direction of this finding is consistent with a recent meta-analysis in which HM-derived fortifiers were associated with lower odds of necrotizing enterocolitis (OR 0.65; 95% CI 0.44 to 0.97), an association that, however, did not persist in the analysis restricted to randomized trials, which are still scarce and small (24).

The most relevant safety finding was the absence of late-onset neonatal sepsis in the LioNeo group, compared with six cases in the HMCA group (*P* = 0.01; PP analysis: *P* = 0.02), a result that should be interpreted with caution because the study was not powered for this outcome and the number of events was small. A plausible hypothesis involves human milk oligosaccharides: in a cohort of Peruvian VLBW infants, oligosaccharide concentrations in maternal colostrum were associated with a lower risk of late-onset neonatal sepsis (HR 0.63; 95% CI 0.41 to 0.97) in an exploratory analysis (25). Meta-analyses of randomized trials and the Swedish multicenter trial by Jensen et al. found no significant reduction in late-onset sepsis with HM-derived fortifiers (5,26,27), and the most recent analysis concluded that the available data are insufficient to confirm or exclude this protective effect, leaving the question open (28).

Serum zinc concentrations decreased similarly in both groups, a pattern consistent with the decline in serum zinc described in the first weeks of life in preterm infants (29) and also observed in the phase 1 trial, which, with 20 participants per group, found no difference in serum copper between groups (30). ESPGHAN recommends an enteral zinc intake of 2 to 3 mg/kg/day for this population, noting that HM alone is not sufficient to meet this recommendation (3). Regarding copper, the HMCA group received more copper (85.4 versus 48.3 µg/100 mL) and about five times more zinc (1.1 versus 0.2 mg/100 mL) but showed a decrease in serum copper, whereas the LioNeo group showed an increase (adjusted difference on day 21: 16.83 µg/dL; 95% CI 11.49 to 22.19). A possible explanation is competition between zinc and copper for intestinal absorption, recognized by ESPGHAN (3). Nevertheless, mean copper concentrations in both groups remained within the reference range described for preterm infants (31).

### Limitations

This study has limitations. The sample size was smaller than planned owing to recruitment difficulties during the COVID-19 pandemic, which reduced the precision of the estimates. For length, however, the confidence intervals remained above the non-inferiority margin in the ITT and PP analyses, with both simple and adjusted models; secondary and safety outcomes should be interpreted as hypothesis-generating. The non-inferiority margin was defined for length based on the statistical variability observed in this population, corresponding to 11% of the expected difference between group means in length, converted to the z-score scale; thus, the clinical relevance of differences of this magnitude has not been established. Although head circumference was prespecified as a primary outcome, no specific margin was defined for it; therefore, the results indicate no difference between groups but do not allow non-inferiority to be claimed. Milk intake, recorded after randomization, was used as a covariate; however, non-inferiority for length was also observed in the simple model. The single-center design and 21-day follow-up limit the generalizability of the findings and do not allow conclusions about long-term outcomes.

### Contributions of the study

To the best of our knowledge, this is one of the first randomized, double-blind trials to evaluate an HM-derived fortifier produced in a public milk bank from donor milk. A meta-analysis highlighted the need for well-designed trials free from industry influence, and the most recent meta-analysis on the topic was funded by a manufacturer of human milk-based fortifier (5,24); in this context, LioNeo provides independent evidence. This model makes it possible to offer an exclusive HM-based diet, recommended by WHO for vulnerable preterm infants, in countries such as Brazil, where the commercialization of human fluids is prohibited (2,6). The results are applicable to hemodynamically stable VLBW infants in neonatal units with access to a human milk bank, a structure available throughout the extensive Brazilian network of human milk banks.

### Conclusions

LioNeo, a fortifier produced from donor HM, was non-inferior to the cow’s milk protein-based fortifier for linear growth in VLBW infants, with no difference in head circumference or weight gain, and no adverse event was more frequent with its use. Multicenter phase 3 trials are needed to confirm and extend these findings.

## ACKNOWLEDGEMENTS

We thank the parents and guardians who entrusted their infants’ participation in this study and the human milk donors, without whom LioNeo would not exist. We also thank the multidisciplinary teams of the neonatal intensive care and intermediate care units of HCFMRP-USP for their dedication and rigor in conducting the protocol, the HCFMRP-USP human milk bank for the production and quality control of LioNeo, and the members of the independent external safety committee for their independent oversight of the study.

## DATA AVAILABILITY

Anonymized individual participant data are not publicly available to protect participant confidentiality but are available from the corresponding author upon reasonable request, including to reviewers during peer review.

## Ethical approval

Research Ethics Committee of HCFMRP-USP (CAAE 96682318.2.0000.5440).

## Trial registration

Brazilian Registry of Clinical Trials (ReBEC), RBR-8nnpfm (UTN U1111-1220-0550), registered on June 4, 2019.

## Contributors’ statement

MCAF conceived and designed the study, drafted the manuscript, contributed to data interpretation, provided critical revisions, and was responsible for milk fractionation and nutritional monitoring of newborns. ACS, MMMP, FC, and LMMF conceived and designed the study, provided critical revisions, and contributed to data interpretation. PPP, TS, AA, and ISD were responsible for data collection and reviewed the manuscript. VSB and LGA were responsible for the Human Milk Bank and fortifier production, and reviewed the manuscript. TMBT was responsible for laboratory analyses and reviewed the manuscript. DCA conceived and designed the study, contributed to data interpretation, provided critical revisions, and performed the statistical analyses. FVU and MMO were responsible for milk fractionation and nutritional monitoring of newborns, and reviewed the manuscript. VNP conceived and designed the study, provided critical revisions, contributed to data interpretation, and coordinated and supervised data collection. JSCJ conceived and designed the study, drafted the manuscript, contributed to data interpretation, provided critical revisions, and approved the final version. All authors reviewed the manuscript, approved the final version as submitted, and agree to be accountable for all aspects of the work.

## Declaration of generative AI and AI-assisted technologies

Use of AI-assisted writing tools is declared in the Methods section.

