## Supplementary File 1 for "LioNeo project: a phase 2, randomized, double-blind, non-inferiority trial of a lyophilized human milk-based fortifier for very low birth weight infants"

### Supplementary File 1. Informed Consent Form (English translation of the model approved by the Research Ethics Committee)

**PATIENT No.: _____**

**INFORMED CONSENT FORM AND CONSENT FOR STORAGE OF BIOLOGICAL MATERIAL**

**Study title:** Nutrition of very low birth weight preterm newborns using a concentrate containing lyophilized human milk – Phase 1 and Phase 2 Study: Safety, tolerability and efficacy

**Principal investigator:** José Simon Camelo Júnior, MD, PhD, Department of Pediatrics, Ribeirão Preto Medical School, University of São Paulo. Avenida Bandeirantes, 3900, 5th floor, room 506, HC Criança, Clinics Hospital of Ribeirão Preto Medical School, University of São Paulo (HCFMRP-USP), Ribeirão Preto, São Paulo, Brazil. Contact phone: +55 (16) 3963-6629.

You are being invited to take part, as a volunteer, in a research study. After being informed about the information below, if you agree to take part in this study, please sign at the end of this document, in two copies. If you do not agree to take part in the study, you will not be penalized in any way, and your care at this Hospital will not be affected.

The aim of the study is to test the efficacy of breast milk concentrated with lyophilizate (powdered breast milk) for feeding very low birth weight newborns (1,500 grams or less), and to compare how the children grow, how they gain weight, head size, among other aspects, and the occurrence of unexpected effects, in comparison with other newborns receiving breast milk enriched with powdered cow's milk.

If you agree to take part, you will allow:

1. First, the review of your child's medical record at HC Criança, to collect birth data (for example, gestational age and type of delivery, what happened during birth, and the conditions at birth);

2. Your child to be randomly assigned to one of the study groups, to receive breast milk with a powdered breast milk additive or breast milk with a powdered cow's milk additive. This assignment is done by computer, which means that no one will know what your child is receiving, except the people responsible for preparing the study milk.

3. Your child to be followed for 21 consecutive days. On every day of follow-up in the study, the following data will be assessed:

    - Body weight and abdominal circumference;
    - Volume of milk ingested;
    - Occurrence of unexpected effects, such as vomiting and regurgitation, changes in stools, abdominal distension, blood in the stools, intestinal perforation and infection.

4. Subsequently, that the following be assessed by the investigator at two different time points:

    - First and last assessment (day 1 and day 21): length, head circumference and blood tests (sodium, potassium, chloride, magnesium, total calcium, phosphorus, copper, zinc, blood glucose, alkaline phosphatase, total protein, albumin, urea, iron, ferritin, hemoglobin and hematocrit), urine and stools.

That two dried blood spot samples on filter paper, urine (for organic acid profile) and stool samples from your child be collected and stored for later analysis. These samples will be kept in a freezer at the Laboratory of Pediatrics, Metals and Rare Diseases for up to 10 years. The investigators responsible for the team undertake to identify the samples and the data collected in a way that guarantees secrecy and confidentiality to you and your child; for this purpose, the blood, urine and stool samples will be identified by numbers (each sample will receive an identification number).

Regarding the material to be stored, a sample of up to 50 milliliters of urine (a small coffee cup), a sample of up to 10 grams of stool (a dessert spoon) and a sample of up to 3.5 milliliters of blood (1 teaspoon) will be collected at each time point (beginning and end of the study). To reduce or relieve any discomfort to your child, the procedures will be performed by a member of the staff who is trained and experienced in this type of collection in the routine assessment of preterm infants. You also have the right to withdraw your consent for the storage and use of the stored biological material at any time.

The risks or discomforts of the study to your child's physical or psychological health are minimal. Blood collection may cause bruising and pain at the puncture site; this risk will be minimized by having the procedure performed by a trained professional. The tests that will be collected are part of the routine assessment of preterm infants. To perform the data collection required for this study, your child will not feel any additional pain or discomfort beyond the routine care procedures performed in the Neonatal Intensive Care and Intermediate Care Units of HC Criança.

The expected benefits of the study are the contribution to knowledge on the efficacy of breast milk concentrated with powdered breast milk for feeding very low birth weight newborns, which may, in the future, improve health care for this population, ensuring equal or better growth and an equal or lower occurrence of adverse effects compared with other newborns fed human milk enriched with a commercial cow's milk-based additive. This study may or may not bring immediate benefits (in growth, weight gain and metabolism) to your child, but it may help improve, in the future, the treatment of very low birth weight preterm patients, and also contribute to a better understanding of growth factors and of the occurrence of adverse effects with the use of human milk concentrated with powdered human milk for feeding these newborns.

Absolute confidentiality of the data obtained individually in this study will be maintained to ensure the privacy of the participants. The investigators will be available to answer any questions at any time during the study, and you are guaranteed access to the results of the study.

This study was reviewed and approved by the Research Ethics Committee (REC) of HCFMRP-USP.

A Research Ethics Committee (REC) is a group of people responsible for overseeing research involving human beings conducted at the Institution, whose role is to protect and guarantee the rights, safety and well-being of all research participants who volunteer to take part in it. The REC of the Clinics Hospital and Ribeirão Preto Medical School is located on the lower ground floor of the hospital and is open Monday to Friday, from 8:00 a.m. to 5:00 p.m., contact phone +55 (16) 3602-2228.

You will receive one copy of this Form, dated and signed, and the other copy will be kept by the research team. By signing this form, you agree to take part in the study.

**Participant information (newborn):**

Name: ____________________________________________________________

Date of birth: ___/___/___

**Legal guardian information (father, mother, both or legal guardian):**

Name: _____________________________________________________________

ID number (RG): ________________________________________________

Taxpayer number (CPF): __________________________________________

Address: __________________________________________, no.__________

City _________________ State ____________

Landline (__) __________ mobile 1 (__) ___________ 2 (__) _____________

Signature: __________________________________ Date: _________________

(Other guardian present)

Signature: __________________________________ Date: ___________________

Name: _____________________________________________________________

Assent signature of mother under 18 years of age:

________________________________________________

Date: ______________________________________________________________

Name: _____________________________________________________________

Below you will also find the contact information of the principal investigator, should you have any questions later.

______________________________ ______________________________

Signature of the collaborating investigator

Name:
ID number:
Date:

Signature of witness in cases of illiteracy

Name:
ID number:
Date:
