## Supplementary File 2 for "LioNeo project: a phase 2, randomized, double-blind, non-inferiority trial of a lyophilized human milk-based fortifier for very low birth weight infants"

### Supplementary File 2. Enteral nutrition protocol for preterm infants

HC Criança - Clinics Hospital of Ribeirão Preto Medical School, University of São Paulo
Department of Pediatrics
**José Simon Camelo Júnior, MD, PhD**

#### Introduction

Feeding preterm infants (< 37 weeks of gestational age) is a nutritional emergency and depends mainly on the infant's clinical condition. Technological advances have allowed increasingly smaller infants to survive, in more severe conditions. These infants do not behave homogeneously, which makes the nutrition of infants with lower gestational age and lower weight even more complex (1).

Ideally, the physiological route, that is, the digestive tract, should be used for feeding whenever possible, but this is not always possible from the outset, and total or partial parenteral nutrition is required.

The milk used initially should be human milk, preferably the mother's own colostrum, but there are limitations that will be discussed below, especially maternal cytomegalovirus (CMV) infection status. When the mother's own milk is not available or is insufficient, pasteurized donor human milk (DHM) from a human milk bank should be used.

Preterm infants can be classified as follows:

a) Low birth weight (LBW) infants: < 2,500 g;
b) Very low birth weight (VLBW) infants: < 1,500 g;
c) Extremely low birth weight (ELBW) infants: < 1,000 g.

#### Goals of nutrition in preterm infants

In 1985, the Committee on Nutrition of the American Academy of Pediatrics (AAP) established that the main goal of nutrition in preterm infants should be to achieve a growth velocity comparable to intrauterine growth at the corresponding corrected gestational age (2). To achieve this goal, the anthropometric parameters of preterm infants should be compared with appropriate intrauterine growth reference curves. For a long time, the Alexander intrauterine growth curve, developed in the United States, was widely used (3). However, subsequent studies reassessed the extrauterine growth of preterm infants, taking into account the physiological constraints and limitations associated with prematurity. These studies showed that the AAP growth goal is often unrealistic in many clinical settings. As a result, new growth curves were developed, including the new Fenton curve (4). The INTERGROWTH-21st International Consortium growth standards are currently recommended (5–7).

It is crucial to emphasize that undernutrition in preterm infants can reduce the number of neurons, with long-term negative effects on behavior, cognitive function, memory and the progression of preexisting or concomitant neurological lesions, such as periventricular leukomalacia and intraventricular/periventricular hemorrhage.

On the other hand, excessively rapid growth may favor harmful nutritional programming, with later development of degenerative diseases, especially metabolic syndrome (obesity, type 2 diabetes, dyslipidemia and hypertension), within the framework known as the developmental origins of health and disease (DOHaD).

Therefore, the goals of nutrition in preterm infants should include growth at an appropriate velocity, mirroring as far as possible what would occur in utero, with awareness of the future impact, good neurological development, minimization of previous impairments and respect for the limitations of prematurity.

**Nutrition for low birth weight infants**

Preterm infants admitted to the neonatal intensive care unit should preferably receive their mother's own colostrum and/or pasteurized colostrum from a human milk bank as early as possible, ideally on the first day of life. Subsequent feeding should continue with the mother's mature milk and/or pasteurized donor human milk. If there are absolute contraindications to breastfeeding, preterm infant formula may be used.

The infant should be hemodynamically stable, with measurable blood pressure or indirect signs of stability, such as good peripheral perfusion, palpable peripheral pulses and normal oxygen saturation (90% to 95%).

Asphyxiated infants, subject to redistribution of blood flow to protect the brain, heart and adrenal glands, are more prone to complications such as necrotizing enterocolitis; therefore, the start of feeding may be delayed at the physician's discretion to between the second and fifth days of life, depending on the severity and consequences of asphyxia.

**Feeding based on gestational age and respiratory status.**

1. If the infant is older than 34 weeks of GA and has no respiratory distress, colostrum may be offered at the proposed initial volume of 20 mL/kg by cup and/or the infant may be put to the breast, provided the mother is able;
2. If the infant is between 32 and 34 weeks of gestational age and has no respiratory distress, the ability to coordinate sucking, swallowing and breathing should be tested (preferably by a speech-language pathologist of the service) before the infant receives milk by cup and/or at the breast;
3. If the infant is < 32 weeks of GA and/or has respiratory distress at birth at any GA (between 32 and 34 weeks of GA and > 34 weeks of GA), regardless of ventilatory support, an orogastric or nasogastric tube should be placed, through which colostrum will be given at an initial volume of 20 mL/kg.

**Monitoring of abdominal circumference**

1. Daily measurement at the level of the umbilicus to establish a reference value;
2. If abdominal distension is suspected (increase of ≥10% in abdominal circumference compared with the last routine measurement considered normal), together with changes in abdominal appearance (red or bluish discoloration, visible peristalsis, tenderness on palpation) and/or vomiting or abnormal stools, a medical assessment should be performed and feeding possibly withheld;
3. Gastric residuals should be checked only in abnormal situations, such as abdominal distension (≥10%), changes in appearance, tenderness, abnormal stools (diarrhea, mucus, pus or blood) or vomiting.

**Milk administration**

1. Milk should be given as syringe boluses, by gavage (using gravity) or by infusion pump;
2. If vomiting and/or abdominal distension occur and are related to the speed of milk administration, infusion pumps may be used for slower administration at the physician's discretion.

**Feeding intervals**

1. For infants weighing more than 1,000 g, initial feeds should be given every 3 hours, advancing as tolerated;

2. A 2-hour interval may be considered based on medical assessment and feeding tolerance;
3. For infants weighing less than 1,000 g, see Table 2 for specific feeding recommendations (9).

Table 1 below presents the proposed feeding progression for infants weighing more than 1,000 g, adapted from the protocol used at Emory University School of Medicine, Atlanta, GA, USA (presentation at the Brazilian Congress of Perinatology) (8). This feeding strategy aims to promote safe and effective weight gain, optimizing nutritional status and minimizing risks such as feeding intolerance and necrotizing enterocolitis.

**Table 1 –** Feeding progression for preterm infants with birth weight above 1,000 g

| Day of enteral nutrition | Volume: mL/kg/day | Feed type | Comments |
|---|---|---|---|
| 1 | 20 | DHM | Trophic feeding; if well tolerated, advance; otherwise, maintain the current volume. |
| 2 | 50 | DHM | Pay close attention to the abdomen, vomiting and the daily measurement of abdominal circumference. |
| 3 | 80 | DHM | Pay close attention to the abdomen, vomiting and the daily measurement of abdominal circumference. |
| 4 | 80 | DHM | Consider starting supplementation with a human milk fortifier (LioNeo or FM85®). |
| 5 | 100 | DHM + fortifier | LioNeo or FM85® – Monitor the abdomen and vomiting closely. |
| 6 | 120 | DHM + fortifier | LioNeo or FM85® – Monitor the abdomen and vomiting closely. |
| 7 | 140 | DHM + fortifier | LioNeo or FM85® – Monitor the abdomen and vomiting closely. |
| 8 | 160 | DHM + fortifier | LioNeo or FM85® – Monitor the abdomen and vomiting closely. |

DHM: pasteurized donor human milk from a human milk bank.

Table 2 below describes the feeding progression for preterm infants with birth weight < 1,000 g, based on the protocol of The Hospital for Sick Children, Toronto, Canada, and affiliated hospitals (9).

**Table 2 –** Feeding progression for preterm infants with birth weight below 1,000 g

| Birth weight | Initial stage (Induction) | Subsequent stage (Progression) | Rate of increase (Thereafter) |
|---|---|---|---|
| 500 to 599 g | 1 mL every 4 h for 3 days | 1 mL every 3 h for 3 days | 1 mL every 2 days |
| 600 to 699 g | 1 mL every 4 h for 3 days | 1 mL every 3 h for 3 days | 1 mL per day |

| | | | |
|---|---|---|---|
| 700 to 799 g | 1 mL every 4 h for 2 days | 1 mL every 3 h for 2 days | 1 mL per day |
| 800 to 899 g | 1 mL every 4 h for 2 days | 1 mL every 3 h for 1 day | 1 mL per day |
| 900 to 1,000 g | 1 mL every 3 h for 2 days | — | 2 mL per day |

**At 100 mL/kg/day – start LioNeo or HMCA fortification**
At 120 mL/kg/day – simplify parenteral nutrition (maintain glucose and electrolyte infusion) and continue LioNeo or HMCA
At 140 mL/kg/day – discontinue parenteral nutrition and continue LioNeo or HMCA
**At 160 mL/kg/day – full-volume feeds, continuing LioNeo or HMCA**

If the infant's weight exceeds 1,000 g, use the criteria in Table 1.

Transpyloric tubes should be placed only if gastric tube placement is unsuccessful, with hard-to-control vomiting and no other clinical findings. Because the transpyloric tube bypasses the gastric barrier, it increases the risk of infection from bacterial translocation and of sepsis. A diagnosis of gastroesophageal reflux disease not controlled with medication may also warrant a transpyloric tube.

Consider the tube to be in the transpyloric position when its tip reaches the angle of Treitz, beyond the fourth portion of the duodenum; take the external crown (level of the anterior fontanelle) to heel measurement, with rapid insertion into the stomach and slower insertion up to the entrance of the jejunum, and confirm the position with X-ray; from this position, always administer the milk slowly by infusion pump.

#### Transition from tube feeding to oral feeding

The transition to oral feeding should take place between 32 and 34 weeks of gestational age, provided the infant is no longer on mechanical ventilation, is receiving full enteral feeding and has undergone a speech-language assessment of the coordination of sucking, swallowing and breathing. If the assessment is adequate, the infant may receive donor human milk by cup and be put to the breast under supervision.

#### Special situations

1. If the preterm infant presents abdominal distension (increase of ≥10% in abdominal circumference compared with the last routine measurement considered normal), gastric residuals may be checked. If they are clear or milky, the volume may be maintained or the previous regimen resumed with a longer interval (from every 3 hours to every 4

hours or even every 6 hours, for example). If they are abnormal, greenish, bloody, dark or fecaloid, feeding should be withheld and the tube left on drainage. If the abdomen is reddish, tense or painful, with or without vomiting, abnormal residuals and/or abnormal stools, feeding should be withheld and necrotizing enterocolitis investigated;

2. Because of the potential risk of vertical CMV transmission, raw milk should be used only above 28 weeks of gestational age at birth or corrected age;
3. Umbilical catheters, CPAP and mechanical ventilation are not contraindications to gastric or transpyloric tube feeding. CPAP may increase the risk of distension due to excess air in the digestive tract;
4. Feeding should be withheld only in the cases listed in the previous paragraph. A venous catheter should not be kept in the hepatic territory, and in these cases feeding is a risk;
5. Infants with repeated episodes of apnea with bradycardia should have the continuation of feeding assessed to avoid an increased risk of bowel hypoperfusion and, consequently, necrotizing enterocolitis;
6. Preterm infants receiving vasoactive drugs should receive gastric or transpyloric feeding only if they are hemodynamically stable;
7. Infants with distension, non-abnormal residuals and frequent vomiting may receive trophic feeding, with a total volume ranging from 10 to 20 mL/kg/day for a predetermined period, usually one week, without daily increases, to stimulate gastrointestinal mucosal trophism and motility, with regularization of the migrating motor complexes;
8. Infants who are able to receive kangaroo mother care should have their sucking-swallowing-breathing coordination assessed and, if it is adequate and gestational age is at or above the 32 to 34-week range, they may be put to the breast and, if necessary, feeding may be supplemented with LioNeo or HMCA.
