## Supplementary File 3 for "LioNeo project: a phase 2, randomized, double-blind, non-inferiority trial of a lyophilized human milk-based fortifier for very low birth weight infants"

### Supplementary File 3. Standard operating manual for portioning of fortified milks

Standard Operating Procedure – SOP

Standard Operating Procedure (SOP) – Nutrition / Portioning / Milk Preparation Room

The nutritionists responsible for portioning (unblinded) will carry out their activities in the milk preparation room of the Clinics Hospital of Ribeirão Preto Medical School (HCFMRP-USP). The nutritionists must complete the preparation steps for entering the room, such as standard hand hygiene and the use of a dedicated gown, mask and cap.

After gowning, the electric water bath must be sanitized daily with 70% alcohol. Next, 6 to 10 liters of purified water from the reverse osmosis system of the milk preparation room must be collected in a sanitized container. If this is not possible, two 5-liter jugs of deionized (or distilled) water must be obtained from the Laboratory of Metals and Rare Diseases of FMRP-USP. This purified water will be used in the water bath for thawing the milk. After the water bath is filled, the chemical thermometer will be placed in its proper position and the equipment will be switched on with the temperature set at 40 °C. The laminar flow hood must also be switched on and, after 30 minutes, sanitized with 70% alcohol according to the milk preparation room's own procedure.

Next, the nutritionists must access REDCap. The enrollment of new participants will be checked daily, together with their allocation to the Delta or Gama group. Randomization will be performed by the statistician in REDCap. The three nutritionists agreed on which group would be Delta and which would be Gama, defining which group will receive human milk with lyophilized human milk (LioNeo group) and which will receive human milk with FM85® (HMCA group). This decision was kept in a sealed envelope inside a locked cabinet. Unblinding will occur only at the end of the project or when the internal or external monitoring committee deems it appropriate.

After each participant is randomized to one of the study groups (Delta or Gama), the nutritionist must access the participants' dietary prescriptions in the HCFMRP-USP system and identify the participants. The nutritionist must record, on the portioning spreadsheet, the volume and frequency of the feeds to be offered to each participant. Based on this information, the nutritionist can then determine the total volume of LioNeo and HMCA to be used in 24 hours, as well as the number of syringes needed for portioning. At this point, the total number of syringes, caps and plastic bags required must be set aside for use.

Once this step is completed, the nutritionist must check the temperature of the freezer and of the water bath by double-checking (equipment display and chemical thermometer). When the water bath reaches 40 °C, the number of bottles needed to prepare the total volume of LioNeo and HMCA must be removed from the freezer. The bottles must then be thawed in the water bath at 40 °C, with frequent shaking of the bottle to speed up thawing and homogenize the product.

**Freezer temperature**

The freezer temperature must be checked twice a day, according to the milk preparation room procedures. The temperature monitor is attached to the side of the freezer, and the log must be signed after each reading. Temperature checks must be performed daily. At the end of each month, the control log must be stored in a safe, locked place.

**Water bath**

Bottles of human milk or LioNeo will be thawed according to the prescription demand. The prescription will be checked, and the bottles containing the amount that best matches the prescribed volume will be thawed to avoid leftovers. Thawing will be performed in the electric water bath located in the milk preparation room and used exclusively by the project. The water bath will be used as described below:

1. Sanitize the water bath with 70% alcohol;
2. Fill it with purified water from the reverse osmosis system or deionized/distilled water, approximately 6 to 10 liters, up to the level of the hot steam outlet inside the equipment;
3. Check the water temperature by double-checking (equipment display and chemical thermometer). The milks must be thawed at 40 °C (±1 °C);
4. Record the temperature daily on the water bath log;
5. After use, the water bath must be cleaned with water and neutral soap and sanitized with alcohol;
6. The jugs used to collect water must be washed once a week with water and neutral soap/detergent, without bleach (requirement of the Health Surveillance Agency).

**Laminar flow hood**

1. Switch on the equipment 30 minutes before starting cleaning;
2. Put on a cap, long-sleeved gown and disposable mask. Gloves are not required;
3. Start sanitizing with a disposable multipurpose cloth soaked in 70% alcohol solution, wiping in a single direction, that is, without back-and-forth movements with the disposable cloth;
4. Clean the corners first;
5. Clean the ceiling, wiping the disposable cloth from the inside outward;
6. Clean the back wall (grid), wiping the cloth from top to bottom;
7. Clean the side walls, wiping the cloth from the inside outward;
8. Clean the base (work surface), wiping the cloth from the inside outward.

While the milk is thawing, and after the laminar flow hood has been sanitized, the nutritionists must place the sterile lining in the hood and open the syringe and cap packages inside the laminar flow hood without touching them. The bottles of LioNeo and pasteurized donor human milk (DHM) must be removed from the water bath as soon as only a small ball of frozen milk remains inside, and placed inside the laminar flow hood. The water bath will then be switched off. Next, the nutritionist must put on sterile gloves and start working. Portioning will be performed in the following order: first the preparation of LioNeo and then of HMCA, when applicable.

**Portioning of LioNeo**

The LioNeo bottles must be opened and handled inside the laminar flow hood. The nutritionist must portion the volume of LioNeo for each infant (according to the medical prescription) into 20 or 60 mL syringes.

**Portioning of HMCA**

The DHM bottles must be opened and handled inside the laminar flow hood. The nutritionist will open the FM85® sachets at the place indicated by the manufacturer. The nutritionist will then add FM85® to the total volume of DHM in the glass bottle, following the standard 4% dilution (one 1 g sachet per 25 mL), as indicated by the manufacturer and in accordance with the standards of the HCFMRP-USP nutrition service. The volume of HMCA for each infant (according to the medical prescription) will then be portioned into 20 or 60 mL syringes.

**Syringe labeling and organization of the milk preparation room**

At the end of portioning, the gloves will be removed and the syringes will be identified with labels containing the recipient's name, the time and the volume to be administered. The labels with the infant's data will be printed and attached to the syringes. The labeled syringes will be stored in plastic bags designated for this purpose (one plastic bag per participant). The bag will be labeled to indicate that it belongs to the project (write LIONEO) and placed in the refrigerator of the milk preparation room until transport to the neonatal intensive care unit or intermediate care unit.

Next, the water bath, already switched off, must be emptied and sanitized with 70% alcohol. The supporting materials used for portioning must be removed from the laminar flow hood and taken to the washing area.

**Transport of milk to the neonatal units**

The labeled syringes, packed in plastic bags, will be removed from the refrigerator of the milk preparation room and taken by the project nutritionists to the neonatal intensive care unit or intermediate care unit in an insulated box with ice packs. In these units, the syringes must be stored on the shelves of the distribution refrigerator, between 5 and 7 °C, and will be administered by the nursing staff within the next 24 hours.

**Receipt and storage of human milk bottles**

Once or twice a week, according to demand, the bottles of frozen milk (DHM and LioNeo) sent by the Human Milk Bank must be received at 1:00 p.m., and the temperature of the insulated box used for transport must be checked.

Next, the DHM and LioNeo bottles must be stored in separate drawers in the freezer dedicated to the project. The freezer temperature must be monitored and recorded daily.

**Data entry in REDCap and in the stock control spreadsheet**

Finally, all handling of LioNeo and HMCA must be recorded in REDCap using the specific form for the milk preparation room. In addition, the nutritionists must record daily, on the stock control spreadsheet, the number of bottles used, as well as their respective registration numbers at the HCFMRP-USP Human Milk Bank.
