## Supplementary File 4 for "LioNeo project: a phase 2, randomized, double-blind, non-inferiority trial of a lyophilized human milk-based fortifier for very low birth weight infants"

### Supplementary File 4. Standard operating manual for data collection

LIONEO

Standard Operating Procedure – SOP

Standard Operating Procedure (SOP) – Data Collection – Nursing

Upon enrollment of the very low birth weight (VLBW) infant in the study, data on maternal and obstetric conditions during prenatal care and birth must be retrieved from the Pregnancy card and/or the electronic medical record, according to the data collection form. The data obtained must be entered into REDCap.

Data collection must take place within the neonatal units participating in the study (intermediate care unit and neonatal intensive care unit), and parents or guardians may remain beside the infant throughout data collection. The assessment environment must be calm, with adequate lighting for the necessary procedures, ensuring the participant's privacy.

All materials used in the project (informed consent forms, sample collection tubes, project notebook, among others) will be stored in a locked cabinet in the intermediate care unit and must be returned to the same place after use (except materials that must be discarded), ensuring the safety and integrity of the study materials and of the professionals involved in the study and employed by the health institution.

On entering the neonatal units, the research nurse must perform hand hygiene with an antiseptic agent (chlorhexidine) and, before touching the incubator or crib and handling the VLBW infant, must perform hand hygiene with 70% alcohol gel.

Study participants will be assessed daily for 21 days, with Day 1 counted from the day on which enteral feeding reaches a volume of 100 mL per kilogram of body weight per day. The research nurse must collect data according to the study flowchart described below:

Figure 1 – Flowchart of participant enrollment and follow-up in Phase 2

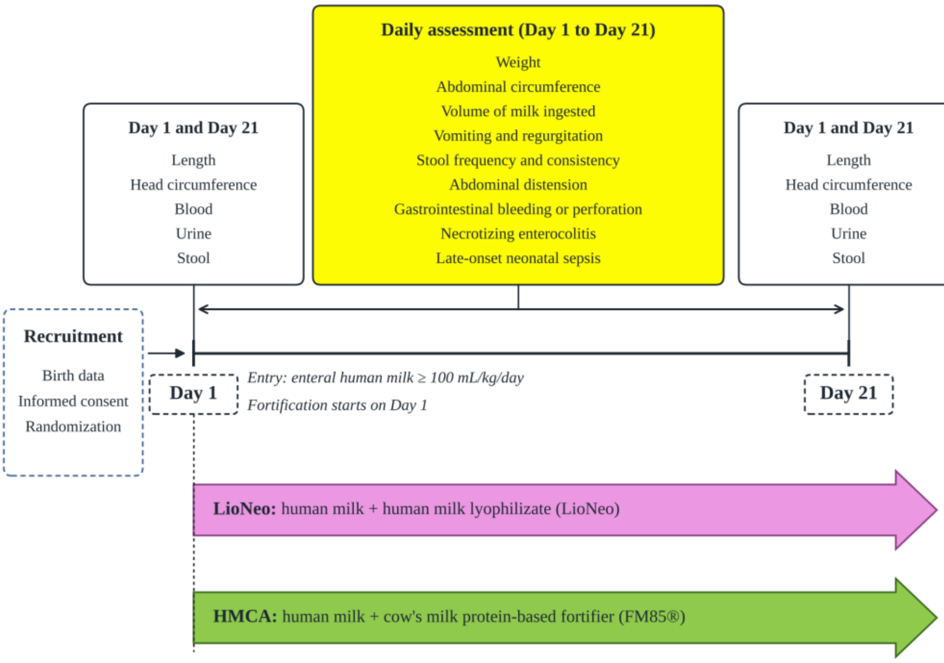


#### Anthropometric data

Anthropometric measurements must be collected in the following order: head circumference, abdominal circumference, length and weight. Measurements must be performed as described below, 2 hours after the last feed or 30 minutes before the next feed.

#### Head circumference

<u>Must be measured and recorded at the infant's enrollment in the study (Day 1) and at the end of follow-up on Day 21.</u>

Head circumference will be measured with a non-elastic, non-stretchable measuring tape with a blank leading section and a precision of 0.1 cm. The research nurse must disinfect the measuring tape with cotton soaked in 70% alcohol before and after use.

First, all accessories on the VLBW infant's head must be removed, as well as any cloth rolls and nests in the incubator or crib that may interfere with the measurement. The research nurse must position the measuring tape with the side marked in centimeters facing outward and the zero mark closest to herself to allow reading.

The measuring tape must be placed over the supraorbital region (front of the head, just above the eyebrows) and pass over the occipital prominence (back of the head, the most prominent posterior point of the head), fitting the participant's head firmly but without compressing it.

The value obtained must be memorized and the head accessories then put back. The VLBW infant must be repositioned in the incubator or crib according to the positioning rotation used by the HC Criança nursing staff. The measured value must then be recorded in the electronic medical record and in REDCap.

**Abdominal circumference**

Must be measured and recorded daily for 21 days, starting from the patient's enrollment in the study.

Abdominal circumference will be measured with a non-elastic, non-stretchable measuring tape with a blank leading section and a precision of 0.1 cm. The research nurse must disinfect the measuring tape with cotton soaked in 70% alcohol before and after use.

For the measurement, the measuring tape must be positioned with the centimeter side facing outward and the zero mark closest to the research nurse, allowing reading. The infant must be placed in the supine position, with the abdomen uncovered and the measuring tape placed at the level of the umbilicus.

The value obtained must be memorized and the VLBW infant repositioned in the incubator or crib, putting back the infant's clothes and any cloth nests or rolls in the incubator or crib, according to the positioning rotation used by the nursing staff. The abdominal circumference value must then be recorded in the electronic medical record and in REDCap.

**Length**

Must be measured and recorded at the infant's enrollment in the study (Day 1) and at the end of follow-up on Day 21.

Length will be measured in centimeters using an anthropometric board made of acrylic plates joined in a rectangular shape, which fits inside the incubator or crib and has a 0.1 cm scale and a minimum length of 0.35 m.

The research nurse must disinfect the anthropometric board by gently rubbing the surface of the device with a cloth moistened with 70% alcohol before each assessment and at the end of data collection.

The VLBW infant must remain in the incubator or crib, be placed in the supine position and be without a diaper or clothes from the waist down; if necessary, perineal hygiene must be performed with cotton and warm water before length is measured. All head accessories, shoes and socks must be removed from the infant, as well as all blankets, nests and/or rolls from the incubator or crib.

Next, the research nurse must hold the infant in one arm and, with the other, place the length board in the incubator or crib and lay the infant in the anatomical position on the base of the board, with the arms resting freely along the trunk, the legs gently extended and the heels together, touching the lower slide of the device. The buttocks, shoulder blades and occiput must touch the horizontal edge of the board.

The research nurse must then move the upper slide until it lightly touches the highest point of the head, applying enough pressure to slightly compress the infant's hair, and the value read must be memorized.

After the measurement, the research nurse must pick up the participant again and remove the device from the crib or incubator. The VLBW infant must be dressed again with clothes, diaper, head accessories, shoes and socks. At the end of the assessment, the research nurse must place the infant in the incubator or crib, following the positioning rotation used by the nursing staff of the HC Criança neonatal units. The length obtained must then be recorded in REDCap and in the electronic medical record.

**Body weight**

Must be measured and recorded daily for 21 days, starting from the infant's enrollment in the study.

Body weight will be measured in grams on an age-appropriate Filizola Baby® electronic scale, calibrated, with a minimum capacity of 125 g and a maximum capacity of 15 kg, a precision of 5 g and a tare of 9 g. To record body weight, the scale must be placed on a flat surface near the VLBW infant's incubator or crib, and the scale pan must be covered with a light, individual cloth. The infant must be placed on the scale in the supine position, without clothes and/or diaper; if necessary, perineal hygiene must be performed with cotton and warm water before the infant is placed on the scale. After placing the participant on the scale, the research nurse must wait for the infant to settle and must remain in front of the scale display until the weight stabilizes and is memorized. The VLBW infant must then be removed from the scale, dressed and repositioned in the incubator or crib, following the positioning rotation used by the nursing staff of the HC Criança neonatal units. The infant's body weight must then be recorded in REDCap and in the electronic medical record.

**Volume of milk ingested**

The volume of milk ingested by the infant must be checked daily in the patient's electronic medical record, from Day 1 to Day 21 of data collection.

The research nurse must check, daily, the feeding prescription written by the HC Criança medical team. To obtain the volume of milk ingested by the VLBW infant, the research nurse will record in REDCap the volume of milk prescribed over 24 hours and the milk leftovers, which will be stored in the neonatal intensive care unit and intermediate care unit in an appropriate container used exclusively by the project so that these leftovers can be checked. The volume will be measured in milliliters, as absolute values and per kilogram of the participant's body weight.

If the infant is on a relactation protocol, the milk delivered in syringes must be offered by cup until the 21 days of follow-up are completed.

**Adverse events**

Adverse events presented by the infant must be checked daily, from Day 1 to Day 21 of data collection, using the HC Criança electronic medical record.

The research nurse must check, daily, the medical and nursing records in the HC Criança electronic medical record, verifying the adverse events presented by the participant. In addition, the research nurse must talk with the nursing and medical staff to ensure that no data are lost. The presence or absence of the adverse events listed below must be checked:

Table 1 – Adverse events checked daily: definition and action

| Event | Definition | Action |
|---|---|---|
| **Vomiting** | Return of a larger amount of food with more complete emptying of the stomach, usually occurring some time after feeding (1). | Check whether vomiting occurred.<br><br>Date and time of onset, frequency, amount and characteristics of the expelled content.<br><br>If it occurs, notify the study medical team and record it in REDCap and in the electronic medical record. |
| **Diarrhea** | Increase in the liquid content of the stools or in the number of bowel movements beyond the normal pattern observed in the infant. | Check whether diarrhea occurred.<br><br>Date and time of onset, frequency, amount and characteristics; stool pattern: (5 = |

| | | |
|---|---|---|
| | | hard; 4 = formed; 3 = soft; 2 = liquid; 1 = watery) (1).<br><br>If it occurs, notify the study medical team and record it in REDCap and in the electronic medical record. |
| **Abdominal distension** | Increase of ≥10% in abdominal circumference compared with the last routine measurement considered normal. | Measure abdominal circumference and, if there is an increase of ≥10% in abdominal circumference compared with the last routine measurement considered normal, pain and/or a reddish abdomen with visible bowel loops, notify the study medical team and the HC Criança medical team. Record the assessment in the HC Criança electronic medical record, together with the names of the physicians notified. |
| **Gastrointestinal bleeding** | Clinical assessment of the occurrence of hematemesis (vomiting of blood), melena (dark stools due to the presence of blood) or hematochezia (presence of fresh blood in the stools). | Check, in the HC Criança electronic medical record and by talking with the staff, whether gastrointestinal bleeding occurred.<br><br>If it occurs, notify the study medical team and enter the data into REDCap. |
| **Necrotizing enterocolitis** | Bell criteria, stage IB or higher, confirmed by clinical and radiological examinations. | Check, in the HC Criança electronic medical record and by talking with the staff, whether necrotizing enterocolitis occurred.<br><br>If it occurs, notify the study medical team and enter the data into REDCap. |
| **Late-onset neonatal sepsis** | Presence of the following signs and symptoms (2):<br><br>General: fever, temperature instability, edema, poor feeding, ill appearance.<br><br>Gastrointestinal: abdominal distension, vomiting, diarrhea or hepatomegaly.<br><br>Respiratory system: apnea, dyspnea, tachypnea, intercostal retractions, nasal flaring, grunting and cyanosis.<br><br>Renal system: oliguria | Check, in the HC Criança electronic medical record and by talking with the staff, whether late-onset neonatal sepsis occurred. If it occurs, notify the study medical team and enter the data into REDCap. |

| | | |
|---|---|---|
| | Cardiovascular system: pallor, mottled, cold or clammy skin, tachycardia, hypotension or bradycardia.<br><br>Central nervous system: irritability, lethargy, tremors, seizures, hyporeflexia, hypotonia, abnormal Moro reflex, irregular breathing, bulging fontanelle, abnormal cry.<br><br>Hematologic system: jaundice, splenomegaly, pallor, petechiae, purpura and bleeding.<br><br>Laboratory: complete blood count with leukocytosis, left shift or leukopenia, with qualitative changes such as toxic granulations and microvacuoles. Identification of bacteria or fungi by blood, cerebrospinal fluid or urine culture.<br><br>Sepsis will be confirmed by culture, and infants will be classified as having suspected or confirmed sepsis according to this criterion. | |
| **Death** | Death from any cause during the study period. For the stopping criterion, an expected mortality below 10% in VLBW infants (≤ 1,500 g) was used as the reference. | In case of death, notify the study medical team and record it in REDCap. |

**Biochemical data**

Venous or arterial blood, stool and urine samples must be collected on Day 1 and Day 21 of data collection to obtain the biochemical data.

Biochemical data will be obtained from venous or arterial blood, stool and urine samples collected by the research nurse on Day 1 and Day 21 of data collection. The results of the biochemical tests must be checked in the HC Criança electronic medical record and entered into REDCap and the electronic medical record. Collection should preferably take place in the morning.

The materials for collecting venous or arterial blood, stool and urine samples must be taken from, and returned when unused to, the place designated for the study materials. The research nurse must set aside the materials needed to collect venous or arterial blood, urine and stool, leaving them close to the infant's incubator or crib.

First, the research nurse must check for stool in the infant's diaper and obtain the sample if the infant has had a bowel movement. Collection will be performed with sterile material (tubes and scoop), and a label must be attached to the tube after it is closed.

Next, perineal hygiene must be performed with cotton and warm water so that the pediatric urine collection bag can be attached. The urine collection bag must be attached only after the research nurse has performed perineal hygiene with cotton and warm water, even if the infant has not had a bowel movement.

The research nurse must then perform venous or arterial blood collection and, at the end, check whether the infant has urinated and/or had a bowel movement in order to collect these samples. At the end of blood, stool and urine sample collection, the infant must be repositioned in the incubator or crib, putting back the infant's clothes and any cloth nests or rolls in the incubator or crib, according to the positioning rotation used by the HC Criança nursing staff.

After all samples from all infants have been collected, labeled and stored in an appropriate container, they must be taken immediately to the laboratory by the project nurse.

**Arterial blood collection**

To perform arterial blood collection, the research nurse must set aside the following materials: one pair of procedure gloves, two cotton balls, 70% alcohol, tray, 3 mL syringe, 25-gauge winged infusion set (butterfly needle), blood sample microtube and the infant's identification label. Pain management during the painful procedure must follow the protocol established by HC Criança (the research nurse must request the pain management protocol from the nurses of the neonatal units), and the necessary materials must be set aside for use. First, the research nurse must check for any condition of the infant that contraindicates the procedure and, if there is none, may proceed with sample collection. The research nurse must perform hand hygiene with an antiseptic agent (chlorhexidine) and, before touching the incubator or crib and handling the infant, must perform hand hygiene with 70% alcohol gel. The research nurse must assess the puncture site, the radial or brachial artery of the right or left upper limb being the sites of first choice. After the puncture site is chosen, the butterfly needle must be connected to the syringe and antisepsis must be performed with cotton soaked in 70% alcohol, using circular movements from the center outward. The butterfly needle must be held bevel up, with the shaft at an angle between 30° and 45°. When the needle is inserted, the research nurse must watch for the return of arterial blood in the butterfly tubing and pull back the syringe plunger until a sufficient amount is obtained for each tube, without exceeding the volume set by international standards as described in the protocol. The needle must then be withdrawn and the insertion site carefully pressed with dry cotton for 5 minutes or until complete hemostasis. At the end, the research nurse must label the microtube with the infant's data and store it in an appropriate container, completing the collection procedure with hand hygiene with an antiseptic agent (chlorhexidine).

**Venous blood collection**

To perform venous blood collection, the research nurse must set aside the following materials: one pair of procedure gloves, two cotton balls, 70% alcohol, tray, 25 × 0.80 mm needle, the infant's identification label and blood sample microtube. The research nurse must perform hand hygiene with an antiseptic agent (chlorhexidine) and, before touching the incubator or crib and handling the infant, must perform hand hygiene with 70% alcohol gel. The research nurse must assess the puncture site, the antecubital fossa and the back of the hand of the right or left upper limb being the sites of first choice. Pain management during the painful procedure must follow the protocol established by HC Criança (the research nurse must request the protocol from the nurses of the neonatal units), and the necessary materials must be set aside for use. After the puncture site is chosen, antisepsis must be performed with cotton soaked in 70% alcohol, using circular movements from the center outward. The needle must be held bevel up, with the shaft at an angle between 15° and 30°. The research nurse must watch for the return of venous blood in the needle hub, positioning the microtube so that blood is collected drop by drop until a sufficient amount is obtained for each tube, without exceeding the volume set by international standards as described in the protocol. The needle must then be withdrawn and the insertion site carefully pressed with dry cotton until complete hemostasis. At the end, the research nurse must label the microtube with the infant's data and store it in an appropriate container, completing the collection procedure with hand hygiene with an antiseptic agent (chlorhexidine).

**Urine sample collection**

To collect a urine sample, the research nurse must prepare: one pair of surgical gloves, one sterile pediatric urine collection bag, one urine sample container, one identification label, one 3 mL or 5 mL syringe, two cotton balls, warm water and a tray. After gathering the materials and performing hand hygiene with an antiseptic (chlorhexidine) and 70% alcohol before touching the incubator/crib or handling the infant, the nurse must check for stool in the diaper. If present, the stool sample must be collected, followed by perineal hygiene with cotton and warm water to position the urine collection bag. Perineal hygiene must be performed even in the absence of stool. In female infants, the urine collection bag must be placed over the labia majora, with the opening covering the urethral area. In male infants, the bag must be positioned so that the penis remains inside the collection bag. The nurse must wait for spontaneous voiding and, during this period, may proceed with blood sample collection. The nurse must then check whether there is urine in the bag. As soon as urine is present, the bag must be carefully removed and perineal hygiene performed with cotton and warm water. The bag may remain attached to the patient until voiding occurs. All the urine collected must be aspirated with a syringe and transferred to the urine sample container, which must be labeled and stored in an appropriate container for transport to the laboratory. The procedure must be completed with hand hygiene using an antiseptic agent (chlorhexidine).

**Stool sample collection**

To perform stool collection, the research nurse must set aside the following materials: one pair of procedure gloves, stool sample container, small spatula, the infant's identification label, two cotton balls, warm water and tray. After gathering the material and performing hand hygiene with an antiseptic agent (chlorhexidine) and 70% alcohol before touching the incubator or crib and handling the infant, the research nurse must check for stool in the infant's diaper and, when present, the sample must be collected and perineal hygiene performed with cotton and warm water. In the absence of stool, the research nurse may proceed with blood collection and check later whether the infant has had a bowel movement. The material must be collected with a small spatula, in the amount of a coffee spoon, and placed in the stool sample container, which must be labeled with the infant's data and stored in an appropriate container to be taken to the laboratory. The procedure must be completed with hand hygiene with an antiseptic agent (chlorhexidine).

All tubes must be wrapped in aluminum foil, and another label must be attached to the outside. These tubes must be transported to the laboratory and stored in a −80 °C freezer. The box number and its storage position in the freezer must be recorded in REDCap.

**Blood pressure**

To measure blood pressure, the research nurse must choose a cuff appropriate for the size of the VLBW infant's limb and connect it to the vital signs monitor, obtaining the values and comparing them with the normal values for birth weight and postnatal age adopted by the service. If the values are within the normal range, the research nurse may consider the VLBW infant a potential candidate for study participation.

**Recording in the source document**

All data collected must be recorded in the patient's electronic medical record in the “Notes” section. When it is not a collection day, record “N/A”. If there are no adverse events, record “None up to the time of collection”.

D (x – record the day and time of the assessment)

Patient assessed according to the LioNeo Project Phase 2 flowchart

Weight:

Abdominal circumference:

Length:

Head circumference:

Blood collection:

Urine and stool collection:

Adverse events:
