## Supplementary Table S1 for "LioNeo project: a phase 2, randomized, double-blind, non-inferiority trial of a lyophilized human milk-based fortifier for very low birth weight infants"

**Supplementary Table S1.** Definitions and severity grading of adverse events

| | | Severity grading | | |
|---|---|---|---|---|
| **Adverse event** | **Definition** | **Mild** | **Moderate** | **Severe** |
| **Diarrhea** | Increase in stool liquid content or in the number of stools beyond the infant's usual pattern, scored as soft (3), liquid (2) or watery (1). | No change in hydration status | – | Profuse |
| **Regurgitation** | Non-forceful return of a small volume of gastric content (milk) to the mouth. | – | – | – |
| **Vomiting** | Return of a larger volume of food with more complete gastric emptying, usually some time after feeding. | Isolated | Repeated, up to 3 times/day | Repeated, more than 3 times/day |
| **Abdominal distension** | Increase of ≥10% in abdominal circumference compared with the last routine measurement considered normal. | 10% to 20% | >20% to 30% | >30% |
| **Gastrointestinal bleeding** | Clinical observation of melena, hematemesis or hematochezia. | Mild | Moderate | Profuse |
| **Gastrointestinal perforation** | Clinical and/or radiological signs of perforation with free air in the peritoneal cavity, after exclusion of necrotizing enterocolitis. | – | – | – |
| **Necrotizing enterocolitis** | Bell criteria stage IB or higher, based on clinical assessment, serial abdominal radiographs and ultrasound, blood counts, C-reactive protein and blood cultures; exploratory laparotomy when indicated. | – | – | Any event |
| **Late-onset neonatal sepsis** | Clinical signs (general, gastrointestinal, respiratory, renal, cardiovascular, neurological or hematological) with laboratory abnormalities (leukocytosis, left shift or leukopenia, toxic granulation). Confirmed when bacteria or fungi were isolated from blood, cerebrospinal fluid or urine culture; otherwise classified as suspected. | Suspected | Confirmed | Suspected or confirmed, with shock |
| **Death** | Death from any cause during the study period. For the stopping criterion, an expected mortality below 10% in VLBW infants (≤1,500 g) was used as reference. | – | – | Any event |

Severity grading refers to event intensity and was defined by the study team before trial initiation; necrotizing enterocolitis, late-onset sepsis and death were considered serious adverse events regardless of severity grade. VLBW: very low birth weight. –: not defined.
