## Supplementary Table S2 for "LioNeo project: a phase 2, randomized, double-blind, non-inferiority trial of a lyophilized human milk-based fortifier for very low birth weight infants"

**Supplementary Table S2.** Incidence of adverse events by group: per-protocol analysis

| Adverse event | LioNeo (n = 61)<br>n (%) | HMCA (n = 55)<br>n (%) | RR (95% CI) | Adjusted RR (95% CI) |
|---|---|---|---|---|
| Diarrhea | 0 (0.00) | 0 (0.00) | –[a] | –[a] |
| Regurgitation | 2 (3.28) | 4 (7.27) | 0.45 (0.08 to 2.36) | 0.45 (0.09 to 2.36) |
| Vomiting | 20 (32.79) | 25 (45.45) | 0.72 (0.45 to 1.14) | 0.78 (0.50 to 1.24) |
| Abdominal distension | 16 (26.23) | 18 (32.73) | 0.80 (0.45 to 1.41) | 0.88 (0.49 to 1.55) |
| Gastrointestinal bleeding | 4 (6.56) | 5 (9.09) | 0.72 (0.20 to 2.55) | 0.74 (0.21 to 2.62) |
| Gastrointestinal perforation | 0 (0.00) | 0 (0.00) | –[a] | –[a] |
| Necrotizing enterocolitis | 1 (1.64) | 3 (5.45) | 0.30 (0.03 to 2.81) | 0.38 (0.04 to 3.47) |
| Late-onset neonatal sepsis | 0 (0.00) | 5 (9.09) | $P = 0.02$[b] | –[a] |

Per-protocol analysis: participants with primary outcome measurements at day 1 and day 21. Incidence: participants with at least one occurrence of the event during the 21 days of study. RR: relative risk (LioNeo vs. HMCA), estimated by log-binomial regression; adjusted RR: adjusted for cumulative milk intake; CI: confidence interval. LioNeo: human milk concentrated with human milk lyophilizate; HMCA: human milk with commercial additive based on cow's milk protein. [a]Not estimable due to zero events in one or both groups. [b]Fisher's exact test.
