## Supplementary Table S3 for "LioNeo project: a phase 2, randomized, double-blind, non-inferiority trial of a lyophilized human milk-based fortifier for very low birth weight infants"

**Supplementary Table S3.** Differences between means of adverse event counts per participant by double Poisson regression: intention-to-treat analysis

| Adverse event | Simple model (95% CI) | Adjusted model (95% CI) |
|---|---|---|
| Diarrhea | –[a] | –[a] |
| Regurgitation | −0.02 (−0.08 to 0.04) | −0.02 (−0.09 to 0.05) |
| Vomiting | −0.20 (−0.50 to 0.10) | −0.28 (−0.82 to 0.26) |
| Abdominal distension | −0.09 (−0.28 to 0.52) | −0.11 (−0.40 to 0.19) |
| Gastrointestinal bleeding | −0.18 (−0.38 to 0.01) | −0.12 (−0.28 to 0.04) |
| Gastrointestinal perforation | –[a] | –[a] |
| Necrotizing enterocolitis | −0.03 (−0.06 to −0.005) | −0.10 (−0.19 to 0.004) |
| Late-onset neonatal sepsis | –[a] | –[a] |

Data are differences between means (LioNeo − HMCA) of adverse event counts per participant during the 21 days of study (95% confidence interval), estimated by double Poisson regression models. Adjusted model: adjusted for cumulative milk intake. LioNeo: human milk concentrated with human milk lyophilizate; HMCA: human milk with commercial additive based on cow's milk protein. [a]Not estimable because the event did not occur in one or both groups.
